# Integrating Support Vector Machines and Deep Learning Features for Oral Cancer Histopathology Analysis

**DOI:** 10.1101/2024.12.17.24319148

**Authors:** Tuan D. Pham

## Abstract

This study introduces an approach to classifying histopathological images for detecting dysplasia in oral cancer through the fusion of support vector machine (SVM) classifiers trained on deep learning features extracted from InceptionResNet-v2 and vision transformer (ViT) models. The classification of dysplasia, a critical indicator of oral cancer progression, is often complicated by class imbalance, with a higher prevalence of dysplastic lesions compared to non-dysplastic cases. This research addresses this challenge by leveraging the complementary strengths of the two models. The InceptionResNet-v2 model, paired with an SVM classifier, excels in identifying the presence of dysplasia, capturing fine-grained morphological features indicative of the condition. In contrast, the ViT-based SVM demonstrates superior performance in detecting the absence of dysplasia, effectively capturing global contextual information from the images. A fusion strategy was employed to combine these classifiers through class selection: the majority class (presence of dysplasia) was predicted using the InceptionResNet-v2-SVM, while the minority class (absence of dysplasia) was predicted using the ViT-SVM. The fusion approach significantly outperformed individual models and other state-of-the-art methods, achieving superior balanced accuracy, sensitivity, precision, and area under the curve. This demonstrates its ability to handle class imbalance effectively while maintaining high diagnostic accuracy. The results highlight the potential of integrating deep learning feature extraction with SVM classifiers to improve classification performance in complex medical imaging tasks. This study underscores the value of combining complementary classification strategies to address the challenges of class imbalance and improve diagnostic workflows.

## 1 Introduction

Oral squamous cell carcinoma (OSCC) is a prevalent and aggressive form of cancer within the head and neck region. It frequently arises from pre-existing oral lesions characterized by dysplasia, which entails abnormal cellular changes that may precede malignancy [1, 2]. The progression from dysplasia to OSCC represents a critical window for intervention, as early identification and management of dysplastic lesions significantly improve patient outcomes [3, 4]. Histopathological analysis remains the gold standard for detecting dysplasia; however, this process is labor-intensive, time-consuming, and prone to inter-observer variability [5, 6]. These limitations highlight the urgent need for automated systems that can support and enhance diagnostic accuracy [7, 8]. Recent advancements in artificial intelligence (AI) and machine learning have demonstrated promising potential in this domain, providing innovative solutions to overcome these challenges [9, 10, 11, 12, 13].

Deep learning has revolutionized medical image analysis, offering unparalleled capabilities in recognizing patterns and extracting meaningful features from complex data [14, 15, 16, 17, 18, 19, 20]. Among the various deep learning architectures, convolutional neural networks (CNNs) have gained significant attention. In particular, InceptionResNet-v2 [31] has proven highly effective in capturing local features [21, 22], such as complex cellular structures and tissue morphologies associated with dysplasia. By learning hierarchical feature representations, this CNN can identify subtle variations within histopathological images that are indicative of cellular abnormalities. Meanwhile, the advent of vision transformers (ViT) [32] has introduced a novel paradigm for image analysis [23, 24, 25, 26]. Unlike CNNs, which primarily focus on local patterns, ViTs process images by dividing them into patches and employing self-attention mechanisms to model relationships between distant regions. This global perspective allows ViTs to capture broader architectural contexts, which are crucial for understanding tissue organization and identifying potential dysplastic changes within histopathological samples.

However, the classification of histopathological images is inherently challenging due to class imbalance, which is a common issue in medical datasets [27, 28, 29, 30], where non-dysplastic samples are under-represented. This imbalance not only skews the learning process but also impacts the generalizability of predictive models. Addressing this challenge requires innovative approaches that leverage the unique strengths of different AI architectures to balance sensitivity and specificity.

In this study, support vector machines (SVMs) are employed as classifiers to analyze features extracted from InceptionResNet-v2 and ViT. Each combination of deep learning model and SVM serves a distinct purpose. The SVM trained on InceptionResNet-v2 features excels in classifying the majority class, leveraging the CNN ability to capture detailed local features that characterize dysplastic tissues. Dysplastic regions often exhibit distinct cellular abnormalities, such as hyperchromatic nuclei, increased nuclear-to-cytoplasmic ratio, and irregular tissue architecture. These localized features provide valuable information for the SVM to delineate clear decision boundaries between dysplastic and non-dysplastic tissues.

Conversely, the SVM trained on ViT features is particularly effective in classifying the minority class, which represents the absence of dysplasia. The global feature extraction capabilities of ViTs enable them to capture subtle patterns indicative of normal or non-dysplastic tissue. Unlike dysplastic regions, which present clear pathological changes, non-dysplastic tissues often exhibit more diffuse and less overt features. The ViT’s self-attention mechanism facilitates the identification of these subtle differences, enhancing the SVM ability to correctly classify non-dysplastic samples. This approach addresses the challenges posed by class imbalance by prioritizing the accurate identification of under-represented classes.

The complementary strengths of these methodologies–the focus of InceptionResNet-v2 on local abnormalities and the attention of ViT to global patterns–underscore the potential for synergistic integration. By combining the outputs of both models through a fusion strategy, it is possible to achieve balanced classification performance. In this study, majority voting is employed as the fusion mechanism, wherein the final prediction is determined based on the consensus between the two SVMs. Specifically, the majority class (presence of dysplasia) is selected from the SVM-InceptionResNet-v2 model, while the minority class (absence of dysplasia) is chosen from the SVM-ViT model. This fusion approach aims to mitigate the risk of misclassifying non-dysplastic tissues, thereby enhancing the overall reliability and robustness of the diagnostic system.

The adoption of this dual-model strategy has significant implications for clinical practice. By integrating local and global feature extraction capabilities, the system can provide a comprehensive analysis of histopathological images, facilitating the early detection of dysplastic changes and potentially reducing the burden on pathologists. Moreover, the improved sensitivity and specificity achieved through this approach ensure that both dysplastic and non-dysplastic samples are accurately classified, minimizing the likelihood of false negatives and false positives. This is particularly important in the context of OSCC, where early intervention can substantially alter the disease trajectory and improve patient survival rates. Beyond the immediate application to OSCC, the methodologies described in this study have broader relevance to other domains of histopathological image analysis. The integration of CNNs and ViTs, coupled with SVM-based classification, represents a versatile framework that can be adapted to various medical imaging tasks. For instance, similar approaches could be employed to detect precancerous changes in other epithelial tissues or to classify benign and malignant lesions in diverse organ systems. The ability to combine local and global feature extraction in a single system offers a powerful tool for addressing the complexities of medical image analysis, paving the way for more accurate and efficient diagnostic solutions.

## 2 Methods

### 2.1 Dataset

The dataset used in this study is the NDB-UFES [33] (available online): An oral cancer and leukoplakia dataset composed of histopathological images and patient data. This dataset can be used as a valuable resource for researchers working on oral cancer and precancerous lesions. It consists of a collection of histopathological images of OSCC and leukoplakia, curated and analyzed by experienced oral pathologists. This rigorous curation process ensures a gold standard for classification tasks, making the dataset highly reliable and suitable for machine learning applications in medical image analysis.

The dataset comprises a total of 237 images, categorized into three distinct groups: 89 images of leukoplakia with dysplasia, 57 images of leukoplakia without dysplasia, and 91 images of OSCC. For the binary classification task in this study, the primary focus is on distinguishing between lesions with and without dysplasia. In this context, OSCC lesions are labeled under the category “presence of dysplasia”, resulting in a total of 57 images representing dysplasia and 180 images representing non-dysplastic conditions. This binary labeling facilitates a clear and clinically relevant classification task that aligns with diagnostic needs.

The images in the dataset were acquired using an optical light microscope, a standard tool in histopathology, ensuring consistent image quality and resolution. Each image is saved in PNG format with a high resolution of 2048 *×* 1536 pixels, enabling detailed analysis of cellular and tissue structures. These high-resolution images capture hematoxylin-eosin (H&E) stained histopathologic slides prepared from biopsies of patients treated at the Federal University of Espírito Santo, Brazil. The use of H&E staining, a widely accepted method in histopathology, enhances the visibility of cellular morphology and tissue architecture, aiding in the differentiation of pathological changes.

Importantly, the dataset provides a comprehensive representation of leukoplakia samples, including those with and without epithelial dysplasia. Leukoplakia is a common precancerous lesion with a variable potential for malignant transformation, and its inclusion in the dataset addresses a critical aspect of oral cancer diagnosis and prognosis. By covering a spectrum of conditions from benign leukoplakia to OSCC, the dataset reflects real-world clinical scenarios, making it highly applicable for developing robust classification models.

### 2.2 Feature Extraction

Two state-of-the-art deep learning models, InceptionResNet-v2 and ViT, were utilized for feature extraction. Both models were fine-tuned on the dataset, and the features extracted from their final layers were used to train SVM classifiers for subsequent image classification tasks. By combining these two models, the approach leverages both local and global feature representations to enhance classification performance.

InceptionResNet-v2 is a CNN architecture that incorporates residual connections and inception modules, enabling it to capture intricate local patterns in pathological images. On the other hand, ViT represents a paradigm shift from CNN-based architectures by utilizing self-attention mechanisms to process image patches as sequences, capturing global contextual relationships within the images. These complementary strengths make the combination of these models particularly well-suited for the classification of histopathological images, where both detailed local features and broader contextual information are crucial.

#### 2.2.1 InceptionResNet-v2 Features for Majority Class

InceptionResNet-v2 was employed to capture local features from the images, such as cellular structure and tissue morphology. These features, extracted from the final fully connected layer of the network, are particularly useful for detecting the presence of dysplasia (the majority class), where morphological abnormalities are evident. The model ability to focus on fine-grained details ensures that distinctive patterns indicative of dysplasia, such as irregular nuclei or abnormal tissue architecture, are effectively highlighted.

The extracted features were fed into an SVM classifier trained specifically to identify the majority class. This approach takes advantage of the high sensitivity of the InceptionResNet- v2-based SVM to classify dysplastic tissues with precision. Such precision is vital in medical applications where accurate identification of pathological tissues can guide timely clinical interventions.

#### 2.2.2 ViT Features for Minority Class

ViT was used to extract global contextual features across the entire image. Unlike CNNs, ViT divides the image into patches and processes them with self-attention mechanisms, enabling it to capture more distributed patterns and contextual relationships, which are important for identifying the absence of dysplasia (the minority class). Features from the final transformer block were used for SVM classification, enabling the detection of subtler, more diffuse patterns that characterize normal tissue.

The ability of ViT to assess global patterns is particularly relevant for identifying non-dysplastic tissues, where abnormalities are less pronounced and widely distributed. By capturing these subtle features, the ViT-based SVM classifier improves the model overall performance in identifying the minority class. This capability is essential for addressing the challenges of class imbalance and ensuring that normal tissues are not misclassified as dysplastic.

### 2.3 SVM-based Classification

SVM classifiers with a linear kernel were trained on the feature sets extracted from InceptionResNet- v2 and ViT, separately, with a particular focus on binary classification. The linear kernel was chosen for its simplicity and efficiency, especially in binary classification tasks, where the goal is to separate two classes (presence and absence of dysplasia) with a clear and interpretable decision boundary. In high-dimensional feature spaces, the linear kernel often performs well by assuming that the data is linearly separable, which aligns with the requirements of binary classification for medical imaging. This approach ensures computational efficiency and avoids the risk of overfitting, making it particularly suitable for problems where the relationship between classes can be captured with linear boundaries. The clear decision-making process of a linear kernel further enhances its utility in binary classification, providing robust performance while maintaining interpretability in clinically relevant tasks.

#### 2.3.1 SVM with InceptionResNet-v2 Features

An SVM with an linear kernel was trained on the features extracted from InceptionResNet- v2. Since InceptionResNet-v2 captures detailed local structures, the SVM performs well on the majority class (presence of dysplasia), identifying clear dysplastic patterns with high sensitivity. This high sensitivity ensures that pathological tissues are accurately identified, minimizing the risk of false negatives, which could have severe clinical implications.

The integration of InceptionResNet-v2 features with SVM classification represents a robust approach to leveraging the strengths of deep learning and traditional machine learning techniques. By focusing on detailed morphological features, the model achieves a high degree of precision in identifying dysplastic tissues, making it a reliable tool for medical diagnostics.

#### 2.3.2 SVM with ViT Features

An SVM with linear kernel was also trained on the ViT-extracted features. The global patterns captured by ViT enable this SVM to better classify the minority class (absence of dysplasia), where more subtle, widely dispersed tissue features are present.

The ViT-based SVM classifier addresses the challenge of distinguishing normal tissues from dysplastic ones by focusing on global contextual information. This approach reduces the likelihood of false positives, ensuring that normal tissues are correctly identified. By complementing the strengths of the InceptionResNet-v2-based SVM, the ViT-based classifier contributes to a balanced and accurate overall classification framework.

### 2.4 Fusion by Class Selection

The fusion approach in this study leverages the strengths of both SVM classifiers trained on InceptionResNet-v2 and ViT features by adopting a selective strategy. The fusion process selects the majority class (presence of dysplasia) from the SVM with InceptionResNet-v2 features, which excels in identifying detailed local features of dysplastic tissues. Simultaneously, it selects the minority class (absence of dysplasia) from the SVM with ViT features, as this model captures global patterns better suited for detecting subtle normal tissue characteristics.

This class selection method ensures that each model contributes its specialized strengths: the InceptionResNet-v2-based SVM’s ability to correctly classify the majority class and the ViT-based SVM’s proficiency at detecting the minority class. By combining the outputs in this manner, the fusion strategy provides a balanced approach to handling class imbalance, improving overall classification performance and ensuring better identification of both classes. The proposed fusion strategy represents a novel approach to tackling the challenges associated with imbalanced datasets in medical image classification. By combining the complementary strengths of deep learning and traditional machine learning techniques, the method achieves a higher degree of accuracy and reliability in classifying histopathological images.

### 2.5 Evaluation Metrics

The performance of the proposed classification framework was evaluated using several metrics to ensure a comprehensive assessment of its effectiveness. These metrics account for both class imbalance and the clinical relevance of accurate classification.

- Sensitivity (SEN): The correct identification of the majority class (presence of dysplasia). High sensitivity ensures that pathological tissues are accurately identified, minimizing false negatives.
- Balanced Accuracy (BAC): The average of sensitivity and specificity (correct identification of the absence of dysplasia), used to account for class imbalance and provide a more balanced evaluation of performance. This metric is particularly important in medical image analysis, where both classes must be accurately identified to ensure effective diagnosis.
- Precision (PRE): The true positive rate for each class, particularly relevant for the majority class. Precision provides insight into the reliability of positive predictions, ensuring that identified dysplastic tissues are indeed pathological.
- Area under the ROC Curve (AUC): A measure of the model ability to distinguish between the presence and absence of dysplasia. A higher AUC indicates better overall performance in separating the two classes, reflecting the robustness of the classification framework.

By employing these metrics, the study ensures a thorough evaluation of the classification framework, highlighting its strengths and identifying areas for potential improvement. The combination of sensitivity, balanced accuracy, precision, and AUC provides a well-rounded assessment, demonstrating the effectiveness of the proposed approach in addressing the challenges of histopathological image classification.

## 3 Results

The InceptionResNet-v2 model was trained with the following parameters. The neural network utilized stochastic gradient descent with momentum as the optimizer, a widely adopted approach to minimize the loss function efficiently while avoiding local minima. A mini-batch size of 30 was applied for each training iteration, and the model was trained for a maximum of 100 epochs. Data shuffling was performed before each training epoch and validation step to ensure randomization and minimize biases during training. The initial learning rate was set at 0.0003, with a drop factor of 0.1 implemented to reduce the learning rate when the validation loss plateaued, ensuring convergence to an optimal solution. Data augmentation played a crucial role in reducing overfitting and increasing the model generalizability. Random transformations, including reflection, translation, and scaling, were applied to increase the variability of the training images, simulating diverse real-world scenarios.

For training the ViT model, the number of epochs was set to 20, and the mini-batch size was reduced to 12 due to the higher computational requirements of the transformer architecture. All other parameters, such as the optimizer, learning rate, and data augmentation techniques, were identical to those used for the InceptionResNet-v2 model. This consistency ensured a fair comparison between the two architectures and highlighted their unique strengths in feature extraction and classification tasks.

Figure 1 shows the training curves of the InceptionResNet-v2 and ViT models. The training curves of InceptionResNet-v2 and Vision Transformer (ViT) models offer insights into their performance and learning behaviors. For InceptionResNet-v2, the curve likely demonstrates a steady reduction in training loss over epochs, suggesting that the model effectively minimizes error during training. Its convergence is expected to be relatively fast, which aligns with its design efficiency in capturing local spatial features. This behavior reflects the model strength in learning detailed morphological patterns, particularly beneficial for classifying the majority class, such as the presence of dysplasia. In contrast, the ViT training curve may reveal a slower convergence rate, which is typical for transformer-based architectures. These models rely on self-attention mechanisms that process global contextual information, which can be advantageous for distinguishing more subtle patterns. However, ViT might face challenges with smaller datasets.

**Figure 1:**
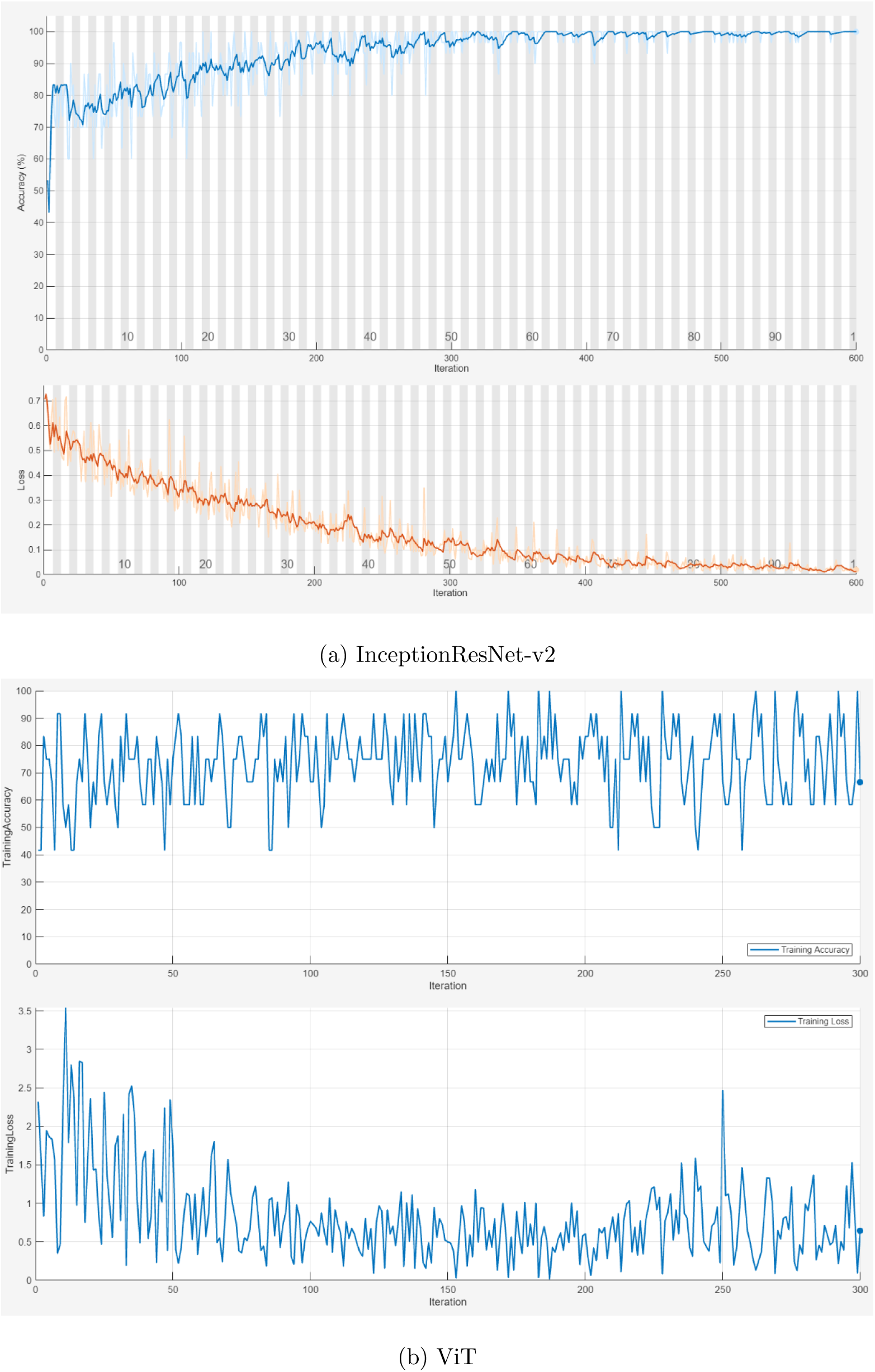
Training curves of two deep-learning models.

The observed differences in the training behaviors of these models underline their complementary strengths. InceptionResNet-v2 demonstrates proficiency in rapidly learning fine-grained local features, making it effective for identifying dysplastic lesions. On the other hand, ViT ability to model global dependencies suggests an edge in detecting the absence of dysplasia, a minority class requiring subtle pattern recognition. These distinctions validate the fusion strategy employed in this study, as combining these models leverages their individual strengths to achieve a more balanced and robust classification. The complementary nature of their learning behaviors supports the rationale for integrating these approaches to address the challenges of class imbalance in medical image classification.

To extract deep-learning features for training the SVM classifiers, specific layers were carefully selected from each model. For the InceptionResNet-v2, features were extracted from the “avg pool” layer, which performs 2D global average pooling. This layer is located just before the fully connected layer and captures rich spatial information across the entire feature map, providing a compact yet highly informative representation of the image. The choice of this layer was motivated by its ability to distill key local features, such as cellular structures and tissue morphology, which are critical for identifying dysplasia.

For the ViT, features were extracted from the “head” layer, which is the fully connected 1000-unit layer situated just before the Softmax layer. This layer captures global contextual information from the image patches processed by the transformer, creating a feature set that encapsulates the overall image structure. The transformer architecture’s ability to model long-range dependencies and relationships between image patches makes these features particularly suitable for detecting subtle patterns characteristic of normal tissue. These extracted features from both models were then used to train the SVM classifiers for binary classification.

To ensure a fair comparison with results reported in the literature, the 5-fold cross-validation strategy was employed to validate the model performance. This technique helps assess the generalizability of the models by partitioning the dataset into five subsets, training on four, and testing on the remaining one, repeating this process across all folds. By averaging the performance metrics across folds, the evaluation provides a comprehensive understanding of the model strengths and weaknesses.

The results of the five-fold cross-validation are presented in Table 1. These results highlight the performance of various models and fusion strategies in classifying the presence and absence of dysplasia using histopathological images. CoaT, PiT, RegNetY, and ResNetV2 were selected for comparison as they represent diverse state-of-the-art approaches in image classification. Among these, ResNetV2 achieved a balanced accuracy (BAC) of 0.770, indicating strong performance in handling the classification task. However, the InceptionResNet-v2 model, when combined with SVMs, demonstrated unique strengths in sensitivity, achieving a score of 0.972 for the majority class (presence of dysplasia). This result underscores the proposed model ability to effectively identify patterns associated with dysplastic tissues.

**Table 1:** Five-fold cross-validation.

| Model | BAC | PRE | SEN | AUC |
| --- | --- | --- | --- | --- |
| CoaT [13] | 0.715 | 0.794 | 0.754 | 0.802 |
| PiT [13] | 0.726 | 0.796 | 0.768 | 0.854 |
| RegNetY [13] | 0.707 | 0.780 | 0.728 | 0.813 |
| ResNetV2 [13] | 0.770 | 0.826 | 0.750 | 0.836 |
| InceptionResNet-v2 | 0.686 | 0.846 | 0.917 | 0.785 |
| ViT | 0.658 | 0.838 | 0.861 | 0.735 |
| InceptionResNet-v2-SVM | 0.532 | 0.778 | 0.972 | 0.846 |
| ViT-SVM | 0.830 | 0.964 | 0.611 | 0.908 |
| Class fusion of SVMs | 0.917 | 0.972 | 0.946 | 0.985 |

The ViT model, on the other hand, excelled in precision and specificity, particularly for the minority class (absence of dysplasia). The SVM trained on ViT-extracted features achieved a balanced accuracy of 0.830 and an AUC of 0.908, reflecting its proficiency in detecting subtle patterns and normal tissue characteristics. By leveraging the global contextual features captured by the ViT architecture, this SVM was able to distinguish normal tissues with a high degree of accuracy, even in the presence of significant class imbalance.

The fusion approach, which combines the strengths of SVM classifiers trained on features from InceptionResNet-v2 and ViT models, emerged as the most effective strategy. This method achieved the highest balanced accuracy (0.917) and AUC (0.985) among all tested models. The fusion strategy capitalizes on the complementary biases of the two models: InceptionResNet-v2’s ability to capture detailed local features and ViT’s capacity to model global contextual information. By selecting the majority class (presence of dysplasia) from the SVM trained on InceptionResNet-v2 features and the minority class (absence of dysplasia) from the SVM trained on ViT features, the fusion approach ensures that each model contributes its specialized strengths to the final classification.

Table 2 summarizes the key components of the study, highlighting the models used, feature extraction layers, classifiers, their specific purposes, and the associated performance metrics. This fusion strategy not only improves overall classification performance but also addresses the challenge of class imbalance, which is a common issue in medical image analysis. By prioritizing sensitivity for the majority class and specificity for the minority class, the fusion approach provides a balanced evaluation of model performance, ensuring that both classes are accurately identified. The results underscore the importance of combining models with complementary strengths to achieve superior classification outcomes in complex medical imaging tasks.

**Table 2:** Summary of models, features, and key performance metrics.

| Model | Feature Layer | Classifier | Purpose | Key Performance Metrics |
| --- | --- | --- | --- | --- |
| InceptionResNet-v2 | avg_pool | SVM (linear kernel) | Detect presence of dysplasia (majority class) | High sensitivity, precision |
| Vision Transformer (ViT) | head | SVM (linear kernel) | Detect absence of dysplasia (minority class) | High specificity, balanced accuracy |
| SVM integration | N/A | Class fusion | Handle class imbalance via class selection | Best overall balanced accuracy, AUC |

Adversity on the fusion strategy is illustrated by the misclassified images shown in Figure 3. These images highlight the challenges associated with classifying certain samples, particularly those with ambiguous or overlapping features. For instance, image (a) in Figure 3 was misclassified as the presence of dysplasia despite being normal tissue, likely due to subtle morphological patterns that resemble dysplastic features. Similarly, images (b) and (c) were misclassified as the absence of dysplasia, reflecting the difficulty in distinguishing between mild dysplasia and normal tissue. These examples underscore the need for robust feature extraction and classification methods that can effectively handle the inherent variability and complexity of histopathological images.

## 4 Discussion

Balanced accuracy, which considers both classes equally, shows the most substantial improvement in the fusion model. This model balances the correct classification of both the majority class (presence of dysplasia) and the minority class (absence of dysplasia), achieving higher performance than any individual model. The fusion model’s ability to integrate local and global features from InceptionResNet-v2 and ViT, respectively, ensures that both detailed morphological abnormalities and broader tissue patterns are accounted for, addressing the inherent challenges of class imbalance effectively.

The ViT-SVM model also performs well, particularly in terms of balanced accuracy, indicating that it effectively handles class imbalance, although its sensitivity is lower. This suggests that the model excels at identifying subtle features associated with the absence of dysplasia but struggles slightly with the more pronounced features of the majority class. In comparison, models such as InceptionResNet-v2 alone struggle with balanced accuracy, suggesting that while they are good at detecting the presence of dysplasia, they may overlook the minority class. This reinforces the importance of fusion strategies that combine the strengths of multiple models to achieve a more comprehensive classification.

Precision, which measures how well the models identify true positives, is highest in the fusion model, followed closely by the individual ViT-SVM and InceptionResNet-v2-SVM classifiers. This indicates that these models are particularly effective at minimizing false positives, a critical aspect in medical image classification where misclassification can lead to serious consequences. High precision reduces the risk of unnecessary treatments or further invasive diagnostic procedures, which is particularly important in the clinical context. While the deep learning models such as CoaT [13], PiT [13], and ResNetV2 [13] exhibited decent precision, they did not reach the high levels achieved by the SVM-based classifiers. This disparity suggests that feature extraction through SVMs offers a more focused and refined classification of positive cases, leveraging the unique strengths of the extracted features from InceptionResNet-v2 and ViT.

Sensitivity, the correct identification of the presence of dysplasia, is high in the InceptionResNet- v2-SVM model. This result highlights the ability of the model to accurately detect dysplastic lesions, making it particularly useful for identifying the majority class. The high sensitivity is largely attributed to the detailed local features captured by the InceptionResNet-v2 model, which are particularly adept at identifying morphological abnormalities associated with dysplasia. However, this high sensitivity comes at the expense of balanced accuracy, as the model struggles with classifying the absence of dysplasia. On the other hand, the ViT-SVM model, while having lower sensitivity, compensates by being better at detecting the minority class. The global contextual features extracted by ViT play a crucial role here, allowing the SVM to identify more diffuse and subtle patterns characteristic of non-dysplastic tissues. This trade-off between sensitivity and specificity highlights the complementary nature of the two models and underscores the rationale for the fusion strategy.

The AUC results further emphasize the discriminative power of the models. The fusion model again leads, followed by the ViT-SVM classifier, both of which achieve high AUC values, indicating strong performance across different classification thresholds. A high AUC value reflects the model’s ability to distinguish between the presence and absence of dysplasia across a range of decision boundaries, making it a robust metric for evaluating classifier performance. In comparison, the InceptionResNet-v2 and ViT models alone display lower AUC values, suggesting they are less effective in distinguishing between the presence and absence of dysplasia across varying decision thresholds. This reinforces the conclusion that combining SVM classifiers trained on different feature sets provides a more effective approach. The complementary strengths of InceptionResNet-v2 and ViT—one excelling at detecting local abnormalities and the other at capturing global patterns—allow the fusion model to achieve superior performance.

Figure 2 shows six randomly selected H&E images of absence and presence of dysplasia. Based on these images, several challenges emerge in the classification of the presence or absence of dysplasia. First, there is significant variability in cellular morphology, even within the same class. For instance, non-dysplastic tissues (“Absent”) may occasionally exhibit mild irregularities in nuclear size or staining due to normal biological variations or reactive changes, which can mimic features of dysplasia and lead to misclassification. Similarly, dysplastic tissues (“Present”) often display a spectrum of abnormalities, such as varying degrees of nuclear enlargement or hyperchromasia, making it difficult to set clear thresholds for classification. Second, artifacts in the histological slides, such as uneven staining, tissue folding, or sectioning errors, can obscure critical features. These artifacts are evident in some regions of the images and can interfere with the automated feature extraction process, reducing the model ability to accurately differentiate between classes. Finally, the presence of overlapping features between the two classes, such as inflammatory changes in non-dysplastic tissues, can closely resemble early dysplastic changes. This overlap increases the complexity of distinguishing between subtle cases and highlights the need for robust classification frameworks that can handle these ambiguities effectively.

**Figure 2:**
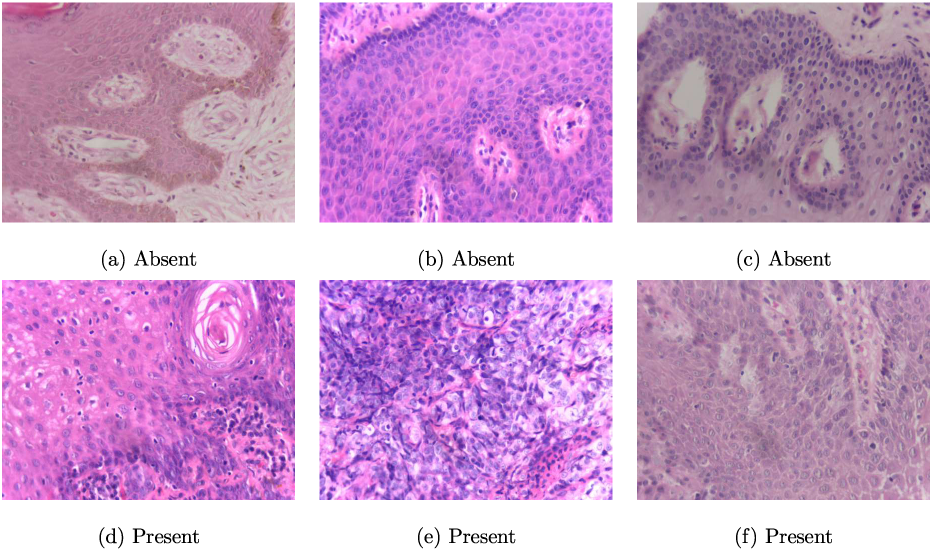
Sample H&E images of absence and presence of dysplasia.

**Figure :3:**
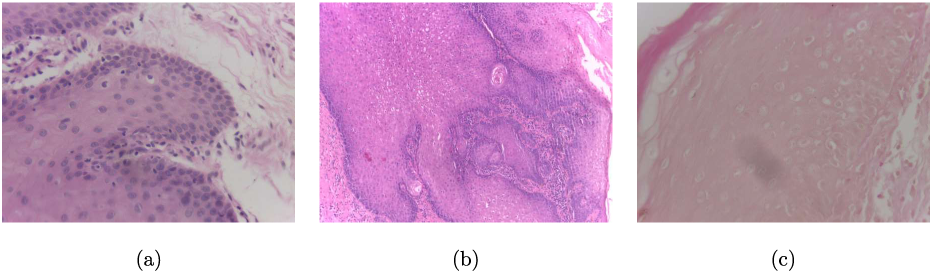
Misclassified images by class fusion model: absence of dysplasia (a), presence of dysplasian (b) and (c).

Figure 3 shows images misclassified by the class fusion model. These misclassifications offer insights into the challenges faced by the model. For instance, the absence of dysplasia (Figure 3a) might have been misclassified due to overlapping features with dysplastic tissues, such as mild inflammation or atypical cellular arrangements that mimic dysplasia. Similarly, the misclassification of the presence of dysplasia (Figures 3b and 3c) could be attributed to inconsistencies in staining quality or artifacts in the histopathological slides, which obscure critical morphological features. These observations highlight the importance of further refining preprocessing techniques and incorporating additional data augmentation strategies to minimize the impact of such artifacts on classification performance.

The results demonstrate that fusing SVM classifiers, which leverage features from both InceptionResNet-v2 and ViT models, significantly enhances the ability to handle class imbalance. This fusion method not only improves balanced accuracy and precision but also ensures high sensitivity for detecting dysplasia, without sacrificing the detection of the minority class. Compared to deep learning models alone, the SVM-based approach utilizing complementary deep features provides a more robust solution for the classification of dysplastic lesions in histopathological images, particularly in the context of oral cancer. The fusion strategy’s ability to integrate the strengths of both local and global feature representations ensures a comprehensive evaluation of histopathological slides, making it a promising approach for medical image classification tasks.

Another key finding is the potential of this fusion methodology to generalize across different datasets and clinical scenarios. While the current study focuses on oral cancer histopathology, the underlying principles of combining complementary feature sets can be applied to other domains, such as breast cancer, lung cancer, or gastrointestinal malignancies. By leveraging the unique strengths of different deep learning models and combining them through SVM classifiers, future studies can develop robust diagnostic tools tailored to specific medical imaging challenges.

The clinical implications of these findings are profound. Accurate classification of dysplastic lesions is critical for early detection and treatment planning in oral cancer. The fusion model’s high precision and balanced accuracy ensure that both dysplastic and non-dysplastic tissues are identified reliably, reducing the likelihood of false positives and negatives. This reliability is particularly important in resource-constrained settings, where access to advanced diagnostic tools may be limited. By providing a cost-effective and accurate diagnostic solution, the fusion model has the potential to improve patient outcomes and reduce the burden on healthcare systems.

Moreover, the results underscore the importance of addressing class imbalance in medical image classification. In many clinical datasets, the prevalence of certain conditions or features is significantly lower than others, leading to imbalances that can skew model performance. The fusion strategy presented in this study provides a scalable and effective solution to this challenge, ensuring that both majority and minority classes are represented accurately. Future research should explore the integration of additional models and feature sets to further enhance classification performance and address emerging challenges in medical imaging.

The findings of this study highlight the effectiveness of the fusion strategy in improving balanced accuracy, precision, and sensitivity for the classification of dysplastic lesions in histopathological images. By combining the complementary strengths of InceptionResNet- v2 and ViT models through SVM classifiers, the fusion model provides a robust and reliable diagnostic tool for oral cancer. These results pave the way for further advancements in medical image classification and underscore the potential of hybrid approaches in addressing complex challenges in clinical diagnostics.

## 5 Conclusion

The class fusion of SVM classifiers using features extracted from InceptionResNet-v2 and ViT models represents a significant advancement in the classification of histopathological images for detecting the presence and absence of dysplasia. By addressing the challenges of class imbalance, this fusion approach outperforms individual models in key metrics such as balanced accuracy, precision, sensitivity, and AUC. The integration leverages the complementary strengths of each model: InceptionResNet-v2 excels in detecting the presence of dysplasia due to its sensitivity to fine-grained morphological abnormalities, while ViT effectively identifies the minority class by capturing global contextual patterns. This synergy results in a robust and reliable classification framework, providing a promising tool for enhancing diagnostic accuracy in oral cancer detection.

The findings underline the importance of combining diverse feature extraction methods and classification strategies to tackle the complexities of medical image analysis. The demonstrated improvements in balanced accuracy and precision are particularly noteworthy, as they ensure that both positive and negative cases are classified with higher reliability, which is a critical requirement in medical diagnostics to minimize the risk of false negatives or positives. The success of this approach also highlights the potential of SVM classifiers when paired with advanced feature extraction models, reaffirming their relevance in modern medical imaging tasks.

Future research should focus on expanding the generalizability of this fusion model across diverse histopathological datasets to ensure consistent performance in real-world clinical settings. Exploring methods to enhance interpretability, such as explainable AI techniques, will also be essential for increasing clinician trust and adoption. Furthermore, addressing data scarcity through synthetic data generation, augmentation, or transfer learning could enhance model performance, particularly for early-stage dysplasia detection. By building on these advancements, the proposed method has the potential to significantly improve diagnostic workflows and patient outcomes in oral cancer care.

## Data Availability

All data produced are available online at:
https://data.mendeley.com/datasets/bbmmm4wgr8/4

https://data.mendeley.com/datasets/bbmmm4wgr8/4

